# Persistent Shift in Laminar Planes Contribute to Post-Surgical Decline in LVEF in Patients with Primary Mitral Regurgitation

**DOI:** 10.1101/2025.04.10.25325619

**Authors:** Louis J. Dell’Italia, Mariame Selma Kane, Jingyi Zheng, Shao-Wei Huang, Betty Pat, Thomas S. Denney

## Abstract

**Background:** The double helical direction of LV laminar sheets from endocardium to epicardium allows for wringing motion or LV twist. This provides a major component to LV wall thickening, stroke volume, and ejection fraction (EF). When this laminar sheet arrangement changes in Primary Mitral Regurgitation (PMR) and whether it reverts to normal after mitral valve repair is unknown.

**Methods:** Normal subjects (n=55) PMR patients had cardiac magnetic resonance imaging (CMR) with tissue tagging and 3-dimensional (3-D) analysis. They were grouped as asymptomatic moderate (n=23) and severe PMR (n=25) by regurgitant volume (RV) and pre-surgery (n=54) with post-surgery follow up at six, 12, and 24 months. Amplitude and directional vector of longitudinal (Ell), circumferential (Ecc), and maximal shortening were computed along with principle strain angles (Ecc°, Ell°, and Err°) at basal, mid, and distal LV levels.

**Results:** Asymptomatic moderate (RV 35 ± 16 ml; LVEF 62 ± 6%) and severe (RV 55 ±16 ml; LVEF 63 ± 6%) and symptomatic pre-surgery (RV 61 ± 29 ml; LVEF 63 ± 8%) had similar increases in mid LV 3-D radius to wall thickness (R/T), decrease in LV mass to volume (M/V) and sphericity index (SI) vs. normal. Radial longitudinal shear strain and mid LV Ecc° and EII° angles increased in all PMR groups, consistent with a shift in LV laminar plane direction and decreased LV SI. Post-surgery, LV end-diastolic (ED) volume, LVED M/V and 3-D R/T returned to normal within two years; however, mid LV circumferential, longitudinal, and maximal shortening decrease below normal. LV Ecc° and Ell° angles, and SI are unchanged from pre-surgery. LVEF decreased post-surgery and had a negative correlation with LV twist at six (r^2^ = 0.30, p < 0.001), 12, (r^2^ = 0.33, p < 0.001) and 24 months (r^2^ = 0.38, p < 0.001) post-surgery.

**Conclusion:** Early changes in Ecc° and Ell° angles, radial longitudinal shear strain, and LV spherical dilatation are consistent with a shift of LV laminar planes that persists after surgery. The extent to which this affects LV twist may underlie a heretofore explanation underlying the decrease in LVEF after surgery for PMR.

## Introduction

Normal left ventricular (LV) geometry is ellipsoidal and becomes spherical as the heart dilates and fails.^1^ We and others have reported that the LV of primary mitral regurgitation (PMR) undergoes spherical remodeling accompanied by an increase in 3-dimensional radius to wall thickness (3D R/T) ratio from base to apex and a decrease in LV end-diastolic (LVED) mass/volume ratio.^2–7^ Dogma of LV eccentric remodeling espouses a series replication of cardiomyocyte sarcomeres as opposed to in parallel in pressure overload.^8^ However, breakdown in the endomysial interstitial collagen scaffolding between cardiomyocytes is an important component in the eccentric remodeling of myofibers sheets in volume overload.^9.10^ This also includes the perimysial collagen connecting myofibers, or bundles of four to six cardiomyocytes, and epimysial collagen connecting myocardial laminae, or myofiber sheets that are oriented in a double helical arrangement from endocardium to epicardium.^11–14^

Fifty years ago Spotnitz and Streeter described extracellular gaps between laminar bundles of myocytes connected by an extracellular matrix.^15,16^ The double helical orientation of these laminar layers from endocardium to epicardial allows for sliding of these laminar layers in a wringing motion that provides a major component to wall thickening, stroke volume and ejection fraction.^1,15,16^ A decrease in LV twist in PMR has been reported with Echo speckle tracking^17–19^ and cardiac magnetic resonance (CMR) tissue tagging in human subjects,^2,6^ and with implanted radiopaque markers in animal studies.^20–23^ In the volume overload model of aortocaval fistula in the dog, Covell and coworkers have demonstrated a time dependent progression of LV spherical remodeling coinciding with change in orientation of myocardial laminae, accounting for the decrease in LV twist in the spherically dilated LV.^24,25^

There has been controversy regarding the mechanism of decrease in LVEF after surgical correction of PMR. Theories have shifted from a mechanical explanation related to a decrease in preload and increase in relative afterload due to correction of the low pressure leak^26,27^ to a more recent myocardial focus related to severe cardiomyocyte ultrastructural damage masked by a normal LVEF^6,28,29^ and increase in extracellular volume with gadolinium enhancement as a surrogate of fibrosis.^30–34^ However, considering its importance to wall thickening, it is unknown whether shifts in myocardial laminar plane orientations return to normal after mitral valve surgery and how it may affect post-operative LVEF. Here we present serial CMR imaging with a 3-D analysis of tissue tagging in which we provide principle strain and shear angles demonstrating the shift of laminar planes in the process of spherical LV remodeling in PMR and its persistence after surgery. The subsequent negative effect on LV twist may explain a heretofore unexplained decrease in LVEF after surgery in PMR.

## Methods

### Patient Population

The study population included 55 normal controls and PMR asymptomatic patients with moderate (n=23) and severe PMR (n=25) grouped by regurgitant volume (RV, moderate 30-40 ml and severe 40-60 ml); and symptomatic (pre-surgery, n=54) with follow up post-surgery at six, 12, and 24 months after mitral valve surgery. Patient recruitment occurred between 2006 and 2010 under National Heart, Lung, and Blood Institute Specialized Centers of Clinically Oriented Research Grant P50HL077100 in cardiac dysfunction. Patients with PMR had echo/Doppler severe isolated mitral regurgitation secondary to degenerative mitral valve disease referred for corrective mitral valve surgery. All pre-surgery patients had cardiac catheterization before surgery and were excluded for obstructive coronary artery disease (>50% stenosis), aortic valve disease, diabetes, or mitral stenosis. Asymptomatic PMR patients with moderate and severe MR had no evidence of coronary artery disease by history or maximal exercise stress test with nuclear perfusion. Normal patients (n = 48) had no prior history of cardiovascular disease or medical illness, no history of smoking, and were not taking any cardiovascular medications. The Institutional Review Boards of the University of Alabama at Birmingham and Auburn University approved the study protocol. All participants gave written informed consent.

### CMR Imaging

Normal subjects and PMR patients underwent CMR on a 1.5-T scanner (Signa, GE) with standard cardiac cine slices in 2- and 4-chamber views, and a short-axis view covering both ventricles and atria. Parameters were set as follows: field-of-view, 360-400mm; 8-mm slice thickness; no gap; and 256*128 matrix. Tagged images were acquired using the same slice prescription as cine with the following parameters: repetition/echo times, 8/4.2 ms; tag spacing, 7 mm; trigger time, 10 ms from the R-wave; and flip angle 10**°**.^5,6,30,31^ In short-axis views, endocardial contours were manually drawn at end-diastole (ED) and end-systole (ES). In all patients, intersections of the mitral and tricuspid valve leaflets with the LV and right ventricular (RV) wall were manually placed in left 2- and 4-chamber view and a right 2-chamber view at ED and ES. All intersections and endocardial contours were propagated to the remaining time frames using an automated algorithm with excellent reproducibility. Mitral regurgitant volume was derived from the difference between LV and RV stroke volumes (SV). The 3-dimensional endocardial circumferential curvature and wall thickness were computed from standard formulas at the wall segments as previously defined in our laboratory.^5,6,30,31^ LA volumes were computed using biplane area length method with manual contours on 2- and 4-chamber long-axis views for each time frame. LA volumetric measurements were provided as maximum atrial volume (Vmax) when the mitral valve opens.

LV twist and shear angle parameters were computed using the Fourier Analysis of Stimulated (FAST) echoes method. 3D LV strains were measured from tagged images at end-systole, which was defined by visual inspection of the image data as the time frame with maximum contraction. Tag lines were tracked with the algorithm described in and edited, if necessary, by an expert user.^37^ The 3D deformation and Lagrangian strain was computed by fitting a B-spline deformation model in prolate spheroidal coordinates to the tag line data.^38^ Normal strains were computed in the radial (Err, maximal shortening), circumferential (Ecc), and longitudinal (EII) directions. Principle strains and associated principal directions and 3D LVES Twist were also computed as described in our laboratory.^39^ All normal, shear, and principal strains were computed at the mid-wall of all segments in the American Heart Association 17-segment model (21) except the apex (segment 17). The first principal strain (E1), which represents the maximum thickening strain, was roughly aligned with the radial direction. The second and third principal strains (E2 and E3) were generally aligned with the longitudinal and circumferential directions, respectively. The third principal strain (E3) corresponds to the maximum contraction strain. Angles between the E3 and circumferential directions (Ecc°), between the E2 and longitudinal directions (Ell°), and between the E1 and radial directions (Err°) were computed. We report circumferential (Ecc), longitudinal (Ell), and maximal (E3) shortening instead of raw strain values (Shortening=strain multiplied by -100%. Shortening is positive and larger values of shortening mean more contraction. For purposes of data analysis, the LV was divided into 3 levels: base, mid, and distal. The strain parameters at each individual level were obtained by averaging the ventricular segments encompassing the whole ventricular wall at that level (6 segments at the base and mid-ventricular levels, 4 segments at the distal level).

### Statistics Analysis

Comparisons among four groups control, moderate, severe and pre surgery (**Figures 1-3**) and pre-surgery with post-surgery follow up at 6, 12 and 24 months, (**Figures 4 and 5**) are tested by Kruskal-Wallis tests, followed by a Benjamini-Krieger-Yekutieli correction. Graphical data are presented as box and whisker plots with median (interquartile range) and minimum and maximum values respectively with each dot representing an individual patient. Spearman correlation was performed between post-surgical LV Twist and LVEF (**Figure 6**). Analyses of repeated measurements across groups are conducted using repeated measures ANOVA and Friedman tests (**Table 1**).

**Figure 1.**
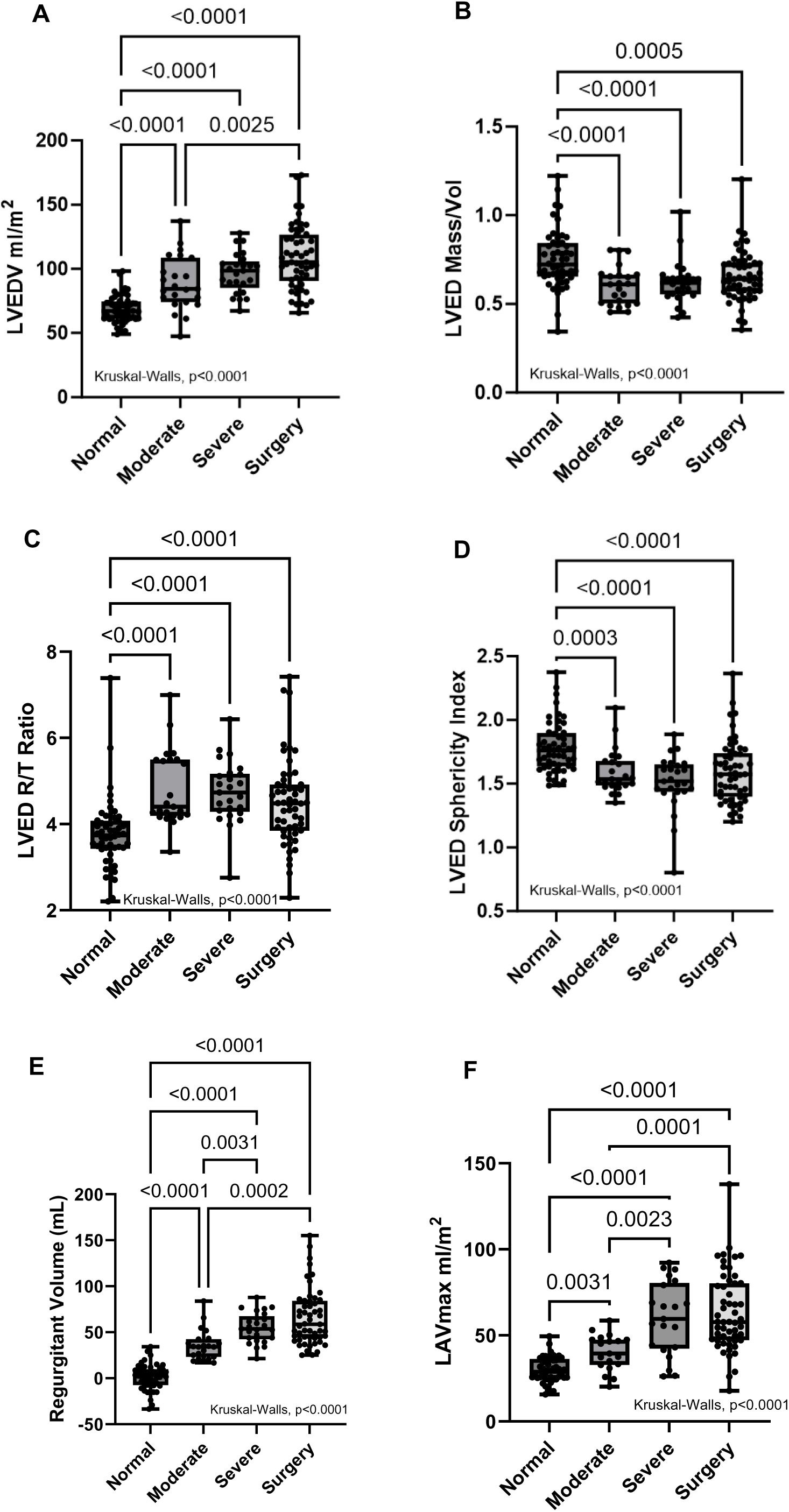
Indices of LV remodeling (A-D) in moderate and severe asymptomatic PMR and pre-surgery PMR. LV end-diastolic volume (LVEDV/m^2^), LVED mass/volume, mid LV 3-D R/T ratio, and LV sphericity index demonstrate similar extremes of eccentric remodeling compared to normals despite an increasing regurgitant volume that is consistent with an increasing left atrial volume (indexed to BSA) (**E and F**).

**Figure 2.**
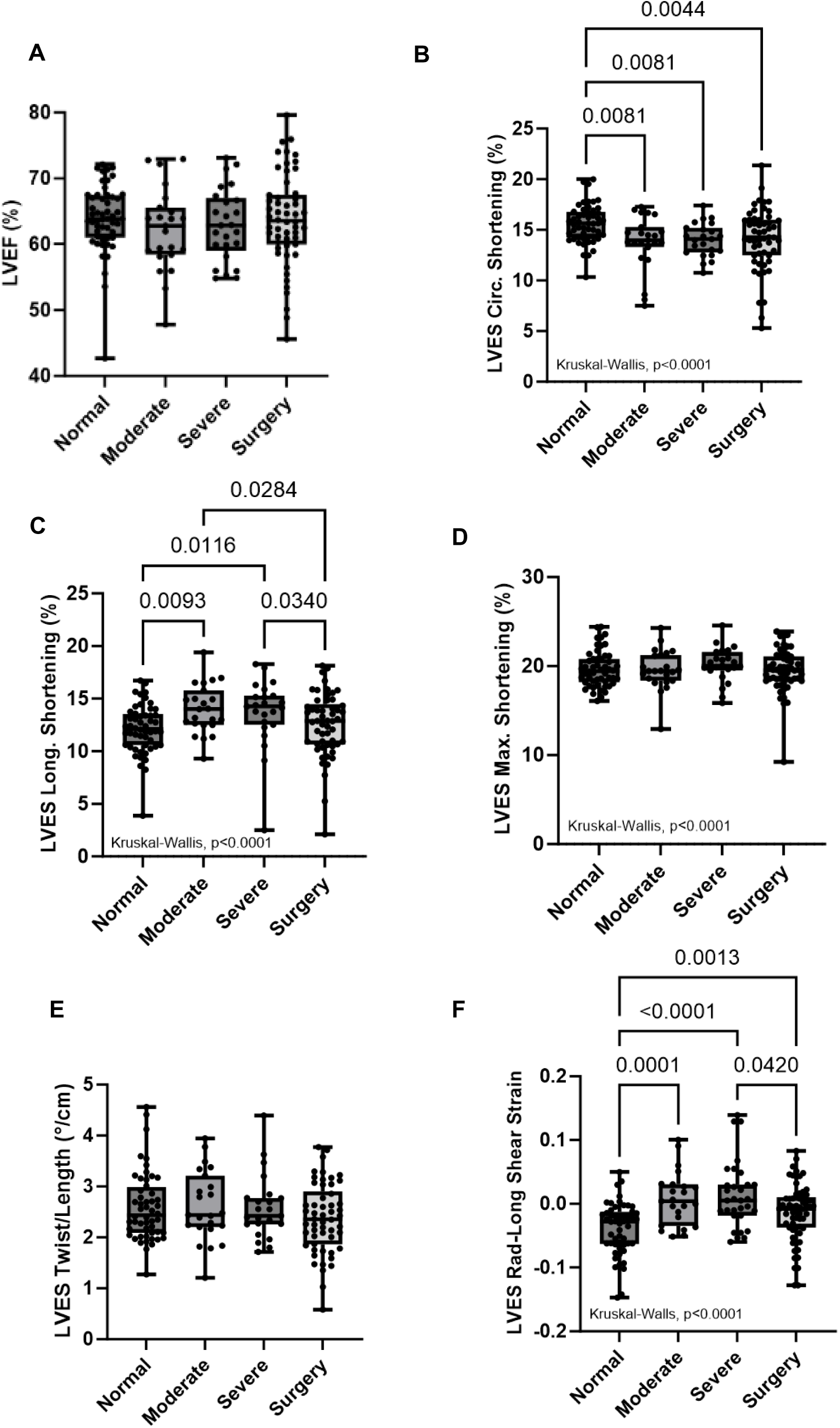
Indices of LV shortening in moderate and severe asymptomatic PMR and pre-surgery PMR. LVEF does not differ from normals (**A**). LVES circumferential (circ.) shortening (**B**) decreases as LVES longitudinal shortening (**C**) increases in asymptomatic moderate and severe PMR; while maximal shortening (**D**) and twist/length (**E**) do not differ from normals. Radial longitudinal shear strain (**F**) increases starting at moderate PMR consistent with a decrease in LV sphericity index (**Figure 1 D**) which is maintained with increasing severity of PMR.

**Figure 3.**
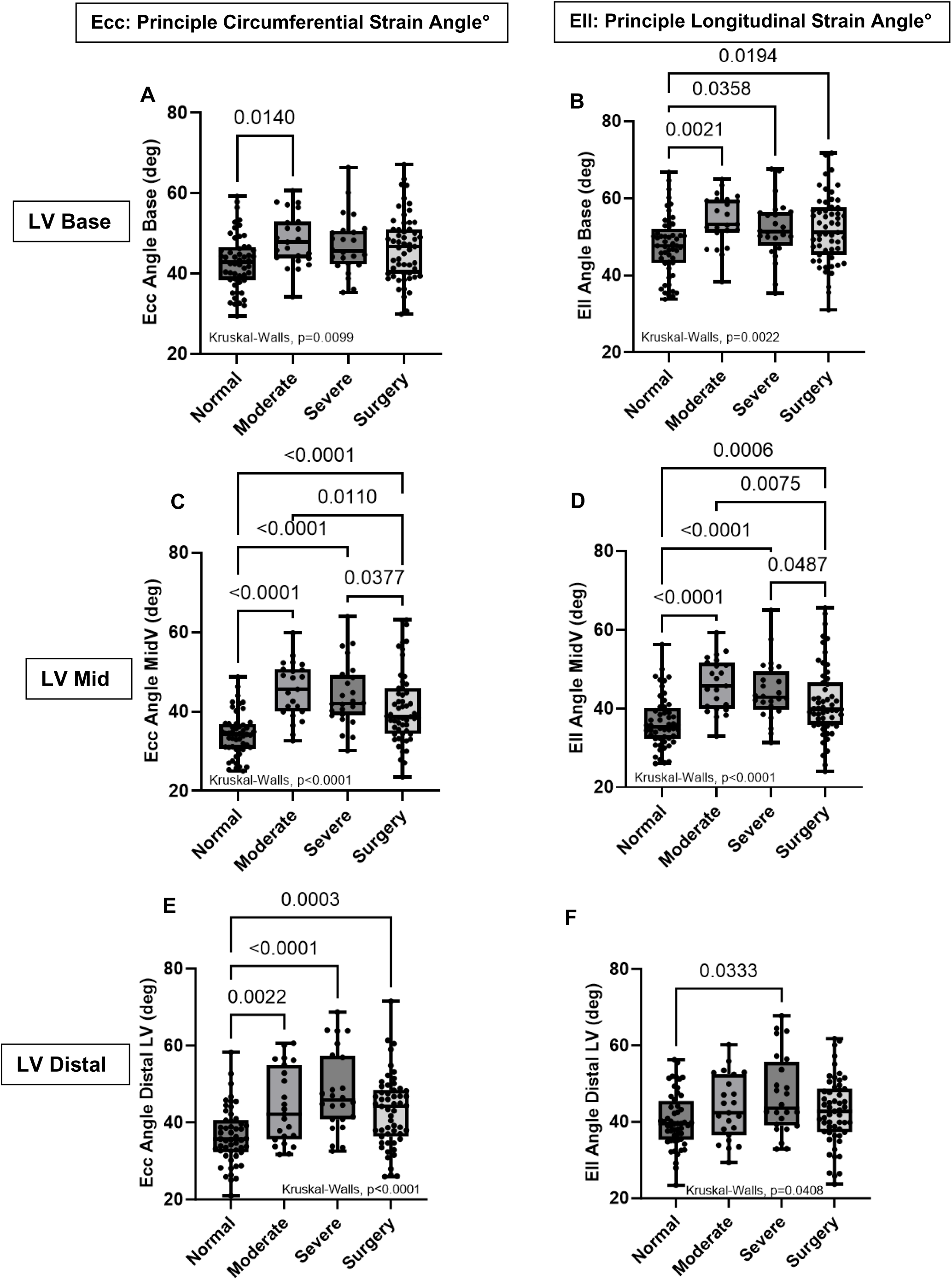
Group Analysis of Principle Circumferential Strain Angle° (Ecc°) and Ell: Principle Longitudinal Strain Angle° (Ell°) at base, mid and distal LV. There were no differences in principle strain and radial angles in PMR vs. Controls (data not shown).

**Figure 4.**
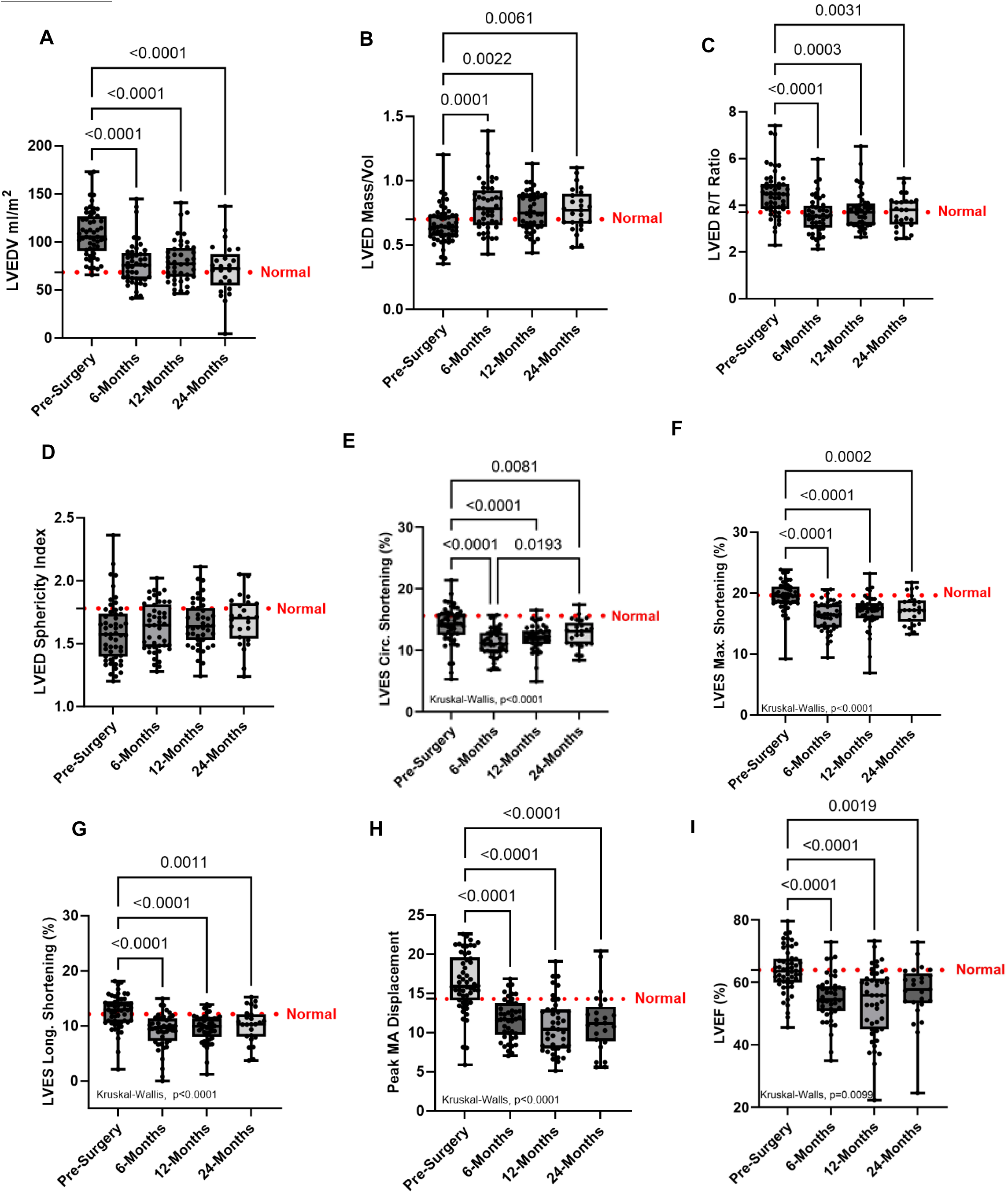
Group analysis of indices of LV remodeling pre- and post-surgery at six, 12, and 24 months (**A-D**). LV end-diastolic volume (LVEDV/m^2^, A), LVED mass/volume (**B**), and mid LV 3-D R/T ratio (**C**) return toward normal while LV sphericity index (**D**) is unchanged from pre-surgery at all post-surgery time points. Post-surgery indices of LV shortening decrease from pre-surgery at six, 12, and 24 months post-surgery (**E-I**).

**Figure 5.**
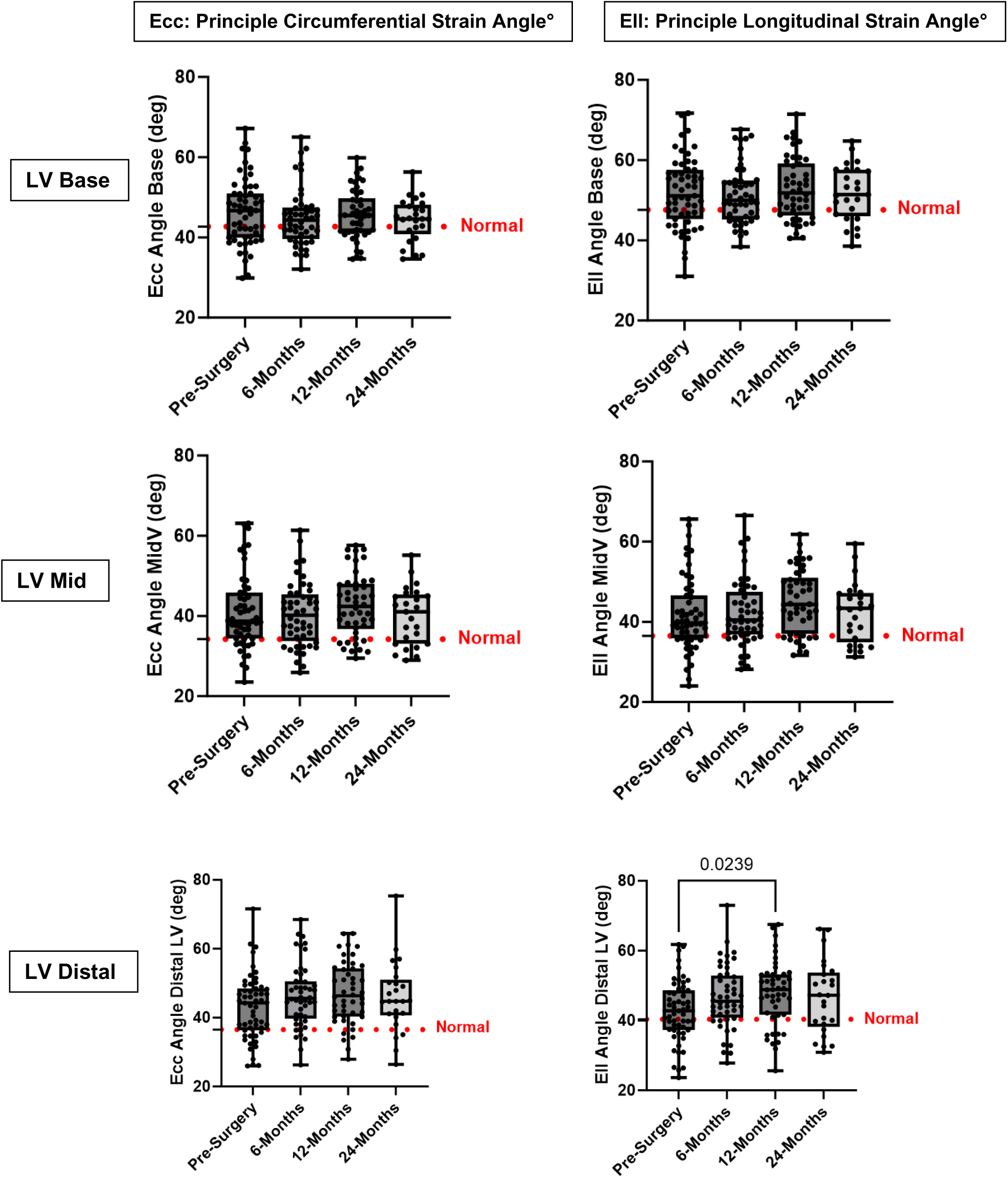
Group Analysis of Principle Circumferential Strain Angle° (Ecc°) and Principle Longitudinal Strain Angle° (Ell°) at base, mid and distal LV post-surgery. Symptomatic PMR patients scheduled for mitral valve repair/replacement surgery (n=39) had follow up every 6 months up to 2 years after surgery with no effect on measured strain angles.

**Figure 6.**
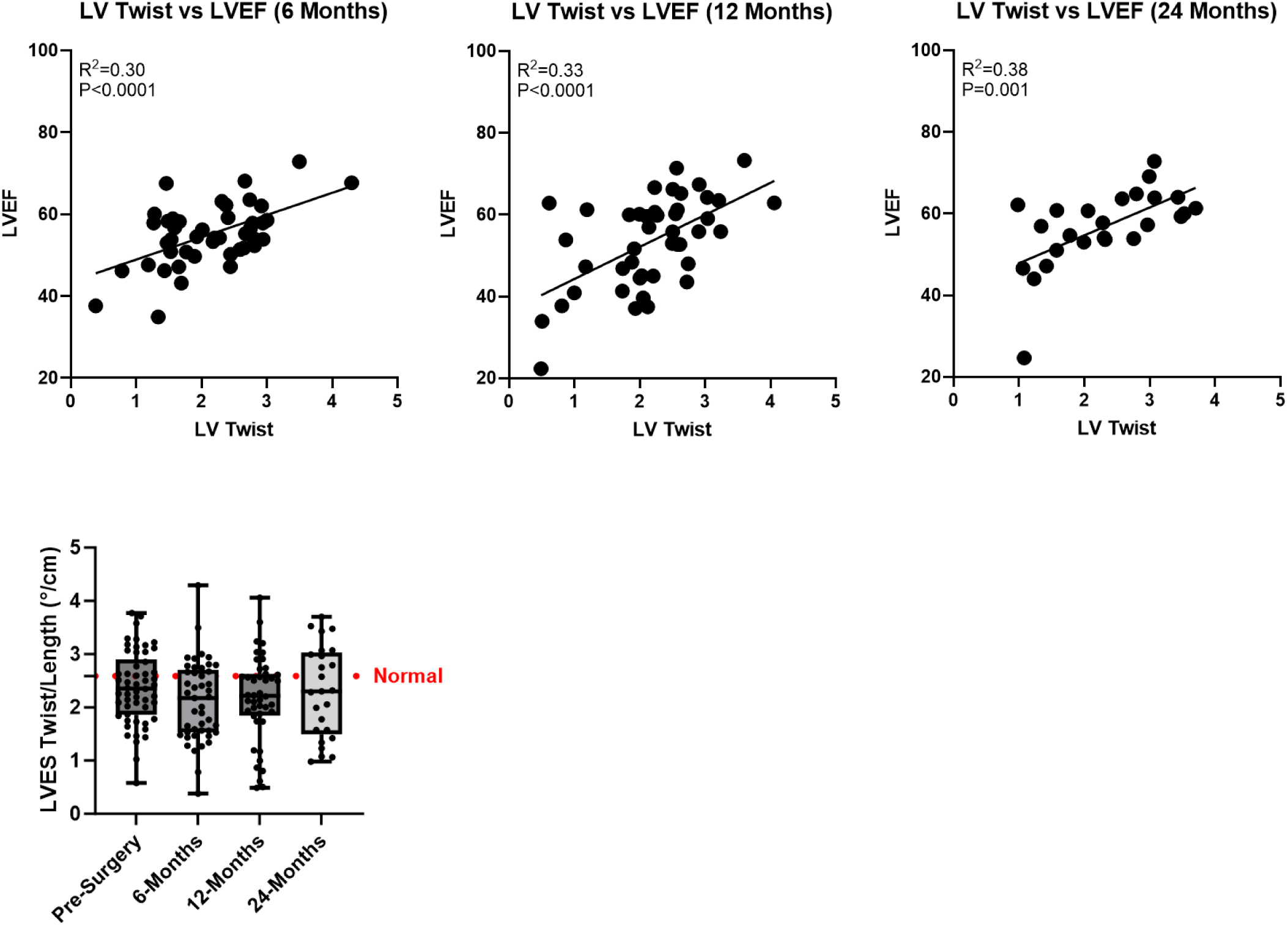
Significant negative relationship between LVEF and LV twist/length in surgical PMR patients at 6, 12, and 24 months post-surgery. LV twist/length did not differ pre and post surgery.

**Table 1.**
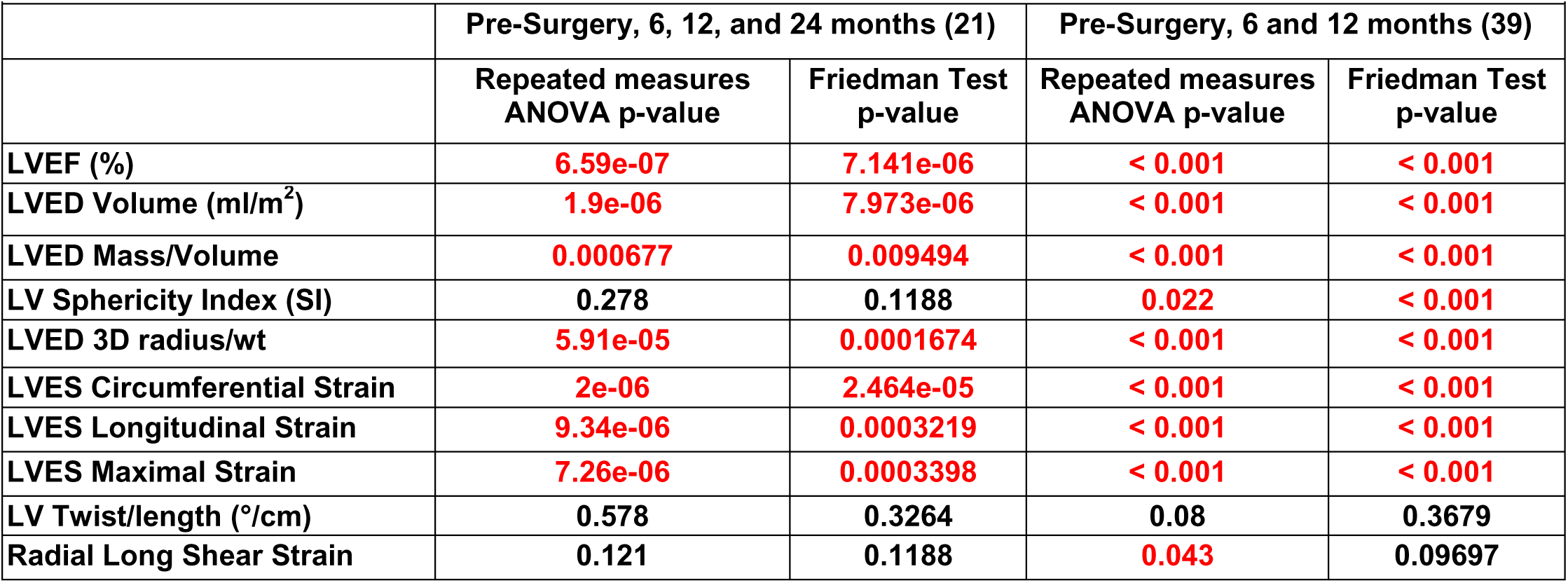
Repeated Measures Analysis for PMR Patients with CMR at Pre-Surgery, 6, 12, and 24 Months (n=21) and for PMR Patients with CMR Pre-Surgery, 6, and 12 months (n=39).

## Results

### Demographics of PMR Patients

Normals were significantly younger (45 ± 15 years; n = 55) than moderate (53 ± 12 years; n = 23), severe (57 ± 9 years, n = 25), and pre-surgery PMR patients (56 ± 12 years, n = 54) (p=0.005). Sex distribution also differed significantly (p=0.03), with normals having a more balanced female-to-male ratio (27:25) compared to the PMR groups.^4,6^ Body surface area was similar across all groups. All moderate and severe PMR patients were asymptomatic (NYHA Class I), with 12 on antihypertensive medications. Pre-surgery PMR patients showed varying symptom levels: 25 Class I, 26 Class II, and 3 Class III. Twenty-two of these patients were taking antihypertensive medications.^4,6^

### Group Analysis of LV Remodeling in PMR versus Controls

Major indices of eccentric LV remodeling including LVED volume, LVED mass/volume, LVED Sphericity Index, and 3-D LVED radius to wall thickness (R/T) are significantly different in normal vs. asymptomatic moderate and severe PMR and pre-surgery PMR (**Figure 1 A-D**). LVEDV is greater in pre-surgery vs. asymptomatic moderate PMR. These indices of LV remodeling do not change in the face of a progressive increase in regurgitant volume that is reflected in increasing left atrial maximal volume (**Figure 1 E and F**). Median LVEF and mid LV maximal shortening (**Figure 2 A and D**) do not differ from normals in all PMR groups; while longitudinal shortening is increased above normal in moderate and severe PMR (**Figure 2 C**) and LV mid circumferential shortening is decreased below normals in all PMR groups (**Figure 2 B**). At the same time, radial longitudinal shear strain increases above normal in all PMR groups while LV twist does not differ from controls (**Figure 2 E and F**). Radial Circumferential and circumferential longitudinal shear strains (**data not shown,**) do not differ from normal in moderate and severe asymptomatic and pre-surgery PMR patients.

There is a significant increase between principle strain and circumferential (Ecc°) and longitudinal (Ell°) angles at the mid LV (**Figure 3 C and D**); while at the LV base Ell° angles are increased at all PMR stages compared to normal (**Figure 3 B**). Left ventricular radial (Err°) strain angles did not differ at LV base, mid wall, and distal in PMR vs normal (**data not shown**). Left ventricular distal circumferential (Ecc°) angles are increased at all PMR stages compared to normal whereas longitudinal (EII°) angle is increased in severe PMR (**Figure 3 E and F**).

### Group Analysis of Post-Surgery LV Remodeling and Function vs. Pre-Surgery

There is a significant decrease in LVEDV and a normalization of the 3-D LVED R/T and LVED mass/volume at six, 12, and 24 months after surgery (**Figure 4 A-C**); however, LV sphericity index is unchanged (**Figure 4 D**). Circumferential (Ecc), longitudinal (Ell), and maximal shortening decrease significantly from pre-surgery. (**Figure 4 E-G**), as well as peak mitral annular displacement and LVEF (**Figure 4 H and I**). Angles between principle strain and circumferential (Ecc°) and longitudinal (Ell°) are unchanged from pre-surgery (**Figure 5**) in the LV base, mid, and distal. Although LV twist did not change post-surgery, there is a wide variability (**Figure 6**), however demonstrates a significant negative correlation between LVEF and LV twist pre-surgery (r^2^ = 0.15, p < 0.004) and post-surgery at 6 months (r^2^ = 0.30, p < 0.001), 12 months (r^2^ = 0.33, p < 0.001), and 24 months (r^2^ = 0.38, p < 0.001).

### Repeated Measures Analysis of LV Remodeling and Function at 6, 12, and 24 Months Post-Surgery (Table 1)

To verify group changes of unequal numbers of patients in **Figures 4 and 5**, we performed repeated measures analysis on 39 surgery PMR patients who had CMR at pre- and post-surgery at six, 12 months, and 21 patients who had CMR at pre-surgery and six, 12 and 24 months post-surgery. Both analyses demonstrate a consistent reverse LV remodeling with a significant decrease in LVEDV and LVED mass/volume and increase in LV 3D radius/wall thickness. Despite reverse remodeling there is a decrease in LVEF and LV circumferential, longitudinal and maximal strains while radial longitudinal shear strain and LV twist do not change from pre-surgical values.

## Discussion

The current study evokes the conundrum that LV remodeling is inherently maladaptive with an early LV spherical transition and decrease in 3-D LV R/T and LVED mass/volume in the low-pressure volume overload of PMR. A decrease in sphericity index parallels a decrease in radial longitudinal shear strain and directional changes in LV principle strain angles in the circumferential and longitudinal direction. These directional changes suggest an early realignment of orientation of laminar planes that persist two years after surgery despite a reversal of eccentric LV remodeling. The extent and persistence after surgery result in a decrease in circumferential, longitudinal, and maximal shortening below normal and a negative correlation between post-surgery LV twist and LVEF for up to two years. Taken together, loss of the normal orientation of the double helical laminar planes may underlie a heretofore-unexplained post-surgery decrease in LVEF.

The current study utilizes a state-of-the-art 3-D analysis that enables calculation of in-plane strains, shear angles, and 3D principle strains occurring in an oblique direction not aligned with any imaging plane. An increase in principal strain angles in the longitudinal and circumferential directions (Ell° and Ecc°) at the mid LV reflects spherical remodeling of the LV. This indicates that LV myocardial fiber sheets are not contracting in their typical aligned pattern especially at the mid LV level and coincides with a significant increase in radial longitudinal shear strain (**Figure 2F**).^38,39^ This occurs at the early moderate stage of PMR and coincides with the decrease in the LV sphericity index. Radial longitudinal shear strain quantifies the interaction or “sliding” between radial thickening and longitudinal shortening. Enhanced radial strain relative to longitudinal strain affects the orientation of the principal strain axes, causing the concomitant increase in radial longitudinal shear strain and increase in Ecc° and Ell° angles. As demonstrated in **Figure 3**, there is a variability across the base to mid to distal LV, consistent with regionally heterogeneous LV remodeling in the dog with volume overload of aortocaval fistula.^24^

While radial longitudinal shear strain is important, global longitudinal strain is widely used clinically and represents the overall shortening of the heart muscle along its length.^40–44^ In the current study, there is an increase in mid LV longitudinal shortening at moderate and severe asymptomatic PMR, while mid LV circumferential shortening decreases at moderate, severe, and pre-surgery stages of PMR. Radial circumferential and circumferential longitudinal shear strains remain normal (data not shown). This is consistent with models that demonstrate the importance of circumferential strain over longitudinal strain in maintaining LVEF in the spherically dilated LV.^45^ Accordingly, we have reported that LV mid circumferential strain rate predicts LVEF below 50% after surgical correction^3^ and signals an incipient decrease in LVEF < 60% in patients with asymptomatic moderate to severe PMR.^4^

To test the validity of the findings in the current study, using the same 3-D analysis with CMR tagging, we demonstrated a directionally opposite decrease in 3-D LV mid and apical principle Ecc° and Ell° angles and an increase in twist in patients with concentric hypertrophy and resistant hypertension.^37^ Thus, the concentric hypertrophic adaptation to pressure overload requires deformation in line with the natural elliptical double helical alignment of laminar planes. In contrast, a reorientation of laminar planes is required to accommodate the low-pressure volume overload of PMR, beyond the limits of individual cardiomyocyte length that begets LV spherical dilatation in PMR. Our findings are consistent with Covell and coworkers who report a time dependent LV spherical remodeling concomitant with a shift of laminar planes using implanted radio-opaque beads in dogs with aortocaval fistula volume overload.^24,25^

The novel finding in the current study is that after surgery, despite a decrease in LVEDV and normalization of LV mid 3-D R/T and LVED mass/volume, there is a persistent 1) increase in principle strain angles (Ecc° and Ell°), 2) decrease in radial longitudinal shear strain, and 3) unchanged sphericity index by group and repeated measures analysis (**Table 1**). The laminar planes are connected by epimysial and perimysial collagen.^11–14^ An increase in extracellular volume by CMR with T1 and gadolinium is an important predictor of outcome in patients with PMR.^30–34^ Although identified as a surrogate of fibrosis, these studies do not include T2 weighting to rule out interstitial edema as a cause of the increase in extracellular volume.^46^ We have reported an increase in interstitial space and extracellular matrix breakdown in both human^6^ and experimentally induced PMR in the dog.^47^ As such, an increase in extracellular volume could also indicate edema and breakdown of epimysial and perimysial structural collagen—a necessary process for realignment of laminar planes.^24,25^ However, development of fibrosis at the level of perimysial and epimysial collagen in a later stage of PMR may prevent realignment of laminar planes after surgery. A recent study reported a small but significant decrease in extracellular volume % following mitral valve repair without a change in late gadolinium enhancement in one third of patients, which may suggest a fibrosis that prevents realignment of laminar planes at a later stage of PMR.^32^

### Limitations

The limited number of patients did not allow for differences between mitral valve repair (n=44) versus replacement (n=10). Although not all patients returned for follow-up CMR, results did not differ for group effects and repeated measures including pre-surgery and post-surgery time points. Using CMR tagging we have assumed that changes in principle strain angles and shear strains reflect changes in the underlying laminar plane architecture and collagen matrix in PMR. Studies using diffusion tensor imaging have demonstrated similar changes in laminar planes in the spherical dilatation of heart failure patients.^48^ Studies using diffusion tensor imaging in PMR will support our novel results in the current investigation, especially in the presence of interstitial edema due to collagen breakdown at earlier stages of PMR.^48^ While the findings of the current investigation are supported in the literature,^38,39^ longitudinal studies are needed to validate the relationship between laminar plane shift and LV remodeling in PMR.

### Conclusions

There is an unmet need for defining optimal timing for surgical intervention in patients with moderate to severe PMR especially in the asymptomatic patient with LVEF > 60%. The current study provides possible evidence for a point of no return of the 3-D laminar structure that is so important in LV twist and its contribution to LV stroke volume and LVEF. Development of symptoms or LVEF < 60% are clearly associated with less favorable outcomes with surgery.^49,50^ There is a need for prospective longitudinal clinical studies that compare the value of strain imaging by Echo speckle tracking or CMR feature tracking or tagging that warn of incipient symptoms or decrease in LVEF < 60% when making clinical decisions for surgical intervention.

## Data Availability

All data will be made available upon request.

## Non-standard Abbreviations and Acronyms

Circ.: circumferential
CMR: cardiac magnetic resonance imaging
EII: longitudinal
Ecc: circumferential
LVED: left ventricular end-diastolic
LVES: left ventricular end-systole
MA: mitral annular
Max.: maximal
M/V: mass to volume
PMR: primary mitral regurgitation
R/T: radius to wall thickness
RV: regurgitant volume
SI: sphericity index

## Acknowledgements

None

